# PyHFO 2.0: An Open-Source Platform for Deep Learning–Based Clinical High-Frequency Oscillations Analysis

**DOI:** 10.1101/2025.05.09.25327327

**Authors:** Yuanyi Ding, Yipeng Zhang, Chenda Duan, Atsuro Daida, Yun Zhang, Sotaro Kanai, Minjian Lu, Shaun Hussain, Richard J. Staba, Hiroki Nariai, Vwani Roychowdhury

**Author notes:** **Correspondence** Vwani Roychowdhury.

## Abstract

Accurate detection and classification of high-frequency oscillations (HFOs) in electroencephalography (EEG) recordings have become increasingly important for identifying epileptogenic zones in patients with drug-resistant epilepsy. However, few open-source platforms offer comprehensive and accessible tools that integrate conventional signal processing with modern deep learning approaches for biomarker analysis. We introduce PyHFO 2.0, an enhanced platform designed for automated detection, classification, and expert annotation of neural events. PyHFO 2.0 includes three commonly used detection methods: short-term energy (STE), the Montreal Neurological Institute (MNI) approach, and a Hilbert transform-based detector. For classification, the platform incorporates deep learning models for artifact rejection, spike-associated HFO (spkHFO) detection, and epileptogenic HFO (eHFO) identification. These models are integrated with the Hugging Face ecosystem for seamless loading and can be replaced with custom-trained alternatives. Furthermore, PyHFO 2.0 features an interactive annotation interface that enables clinicians and researchers to inspect, verify, and refine automated results. The platform was validated using clinical EEG datasets from both human and rodent models of epilepsy, confirming its reliability. PyHFO 2.0 aims to simplify the use of computational neuroscience tools in both research and clinical environments by combining methodological rigor with a user-friendly graphical interface. Its scalable architecture and model integration capabilities support a range of applications in biomarker discovery, epilepsy diagnostics, and clinical decision support, bridging advanced computation and practical usability.

## 1 INTRODUCTION

High-frequency oscillations (HFO) in intracranial electroencephalography (EEG) have been increasingly recognized as a promising biomarker for the localization of epileptogenic zones in human and animal studies (Weiss et al. 2018, Boran et al. 2019, Dimakopoulos et al. 2021, Zweiphenning et al. 2022, Jacobs et al. 2018). A growing body of evidence suggests that the resecting of HFO-generating regions correlates with improved postoperative seizure outcomes, highlighting the clinical relevance of accurate HFO detection and classification (Akiyama et al. 2011, Wu et al. 2010, Jacobs et al. 2010, van’t Klooster et al. 2017, Monsoor et al. 2023, Zhang et al. 2022a Foffani et al. 2007). However, automatic HFO detection is prone to false positives, and distinguishing clinically relevant events from noise or physiological activity often requires both computational model assistance and interactive visualization tools to support expert review.

During the past decade, multiple software tools have been developed to assist in HFO detection, visualization, and analysis. Among them, RIPPLELAB (Navarrete et al. 2016) stands out as an influential opensource MATLAB-based application that consolidated the mainstream HFO detection algorithms along with basic EEG visualization features. The accessibility of RIPPLELAB allowed many studies to standardize

HFO detection methods (Nariai et al. 2020, Gliske et al. 2018, Kuroda et al. 2021, Lisgaras and Scharfman 2023, Barth et al. 2023, Petito et al. 2022), accelerating the adoption of HFO research in various clinical and research settings. Concurrently, multiple open-source EEG processing frameworks-such as MNE (Gramfort et al. 2013), YASA (Vallat and Walker 2021), and PyEEG (Bao et al. 2011) - emerged to support various neurophysiological biomarkers. However, despite recent advances in deep learning for HFO classification, existing open-source frameworks lag behind in integrating these techniques into practical applications.

To bridge the gap between these evolving methods and clinical utility, our previous work introduced a software platform **PyHFO** (Zhang et al. 2024b) as a foundational step towards integrating epilepsy research, particularly HFO studies, with advanced signal processing and deep learning techniques. However, the initial version had several limitations. It lacked a user-friendly interface and did not support integration between model predictions and expert review or annotation. While the platform offered a scalable architecture, it provided only a limited selection of detectors and classifiers. A truly effective system requires the ability to incorporate new detection methods, classification models, or biomarkers to reflect the heterogeneous and evolving landscape of HFO research. Ideally, such a platform should also integrate seamlessly with modern machine learning ecosystems, enabling researchers to deploy state-of-the-art tools with minimal technical overhead.

In this paper, we present **PyHFO 2.0**, a significantly upgraded version of our original platform to further improve clinical engagement by adding: (1) an annotation window that enables in-depth review, verification, and manual annotation of detected HFO events; (2) a pipeline enabling users to train or fine-tune their own deep learning classifiers, facilitate user-driven model customization, incorporating models hosted on Hugging Face for automatic downloading and inference; (3) demonstration of such pipeline by incorporating a state-of-the-art eHFO classifier into the ecosystem; (4) further enhance funcionalities of PyHFO by adding a new Hilbert-based detector and adding user interface (UI) designed for greater intuitiveness and expanded scalability. These new features underscore the growing emphasis on flexible, userfriendly software that can adapt rapidly to both clinical and research demands. By integrating with the Hugging Face ecosystem, PyHFO 2.0 also establishes a direct link to the broader machine learning community, encouraging faster adoption of cutting-edge models for EEG analysis.

The remaining sections of this paper are organized as follows. First, we outline the overall workflow of PyHFO 2.0. We then detail each supported functionality. Next, we present the validation and benchmarking results for these methods, demonstrating their performance across diverse datasets. Finally, we discuss potential extensions—such as cursor support for signal inspection, broader biomarker integration, and the adoption of newer deep learning techniques—and conclude by considering how PyHFO 2.0 can further accelerate research and clinical translation in epilepsy care.

## 2 METHODS AND MATERIALS

### 2.1 Data Acquisition

EEG data used for software development, model training, and evaluation were drawn from three datasets previously described in our earlier work (Zhang et al. 2024b).

#### UCLA iEEG Dataset

Intracranial EEG recordings from 19 patients (10 female, 9 male; ages 3–20 years) with drug-resistant focal epilepsy, each consisting of a 10-minute segment during slow-wave sleep. Data were acquired using grid/strip electrodes at a 2,000 Hz sampling rate (Nariai et al. 2019, Zhang et al. 2022b).

#### Zurich iEEG HFO Dataset

Five-minute interictal sleep EEG segments from 20 patients (16 female, 14 male; age range: 17–52 years), recorded with both grid/strip and depth electrodes, originally sampled at 4,000 Hz and downsampled to 2,000 Hz. Bipolar referencing followed the procedure in (Fedele et al. 2017).

#### UCLA Rodent Dataset

Intracranial EEG (iEEG) recordings were obtained from two adult male Sprague-Dawley rats (300–350g) using depth electrodes targeting the neocortex and hippocampus. One animal received a traumatic brain injury, while the other served as a sham control. (Santana-Gomez et al. 2019).

### 2.2 Requirements

The PyHFO 2.0 software is a multi-window GUI developed in PyQt. It is intended to be a user-friendly and intuitive tool that users with technical and non-technical backgrounds can use to detect and classify HFOs in a time-efficient way. PyHFO 2.0 has been released under Academic Licenses (Licenses for Sharing Software Code Non-commercially, UCLA TDG). The GUI interface was implemented in PyQt version 5.15 to make it compatible with hardware platforms such as Mac OS, Linux, and Windows. The HFO detectors were implemented in Python 3.9.0, and the DL-based classifier was implemented in PyTorch 2.0 and Transformers 4.49.0. We chose Python as the programming platform because it is widely used in large-scale data analysis and deep learning in the medical image field.

### 2.3 Code Availability

The open-source software package is freely available on GitHub (https://github.com/roychowdhuryresearch/pyHFO/tree/pyBrain), and the deep learning model checkpoints used for classification are hosted on Hugging Face (https://huggingface.co/roychowdhuryresearch)

### 2.4 Getting Started

To help users become familiar with PyHFO 2.0, we provide a set of instructional videos that walk through the graphical interface, highlight PyHFO 2.0: An Open-Source Platform for Deep Learning–Based Clinical High-Frequency Oscillations Analysis key features, and demonstrate how to process and analyze EEG data step by step. For users interested in customizing the deep learning component, two example scripts are included. One script shows how to train a new classification model using the platform’s built-in data format, while the other provides a way to integrate an existing pre-trained model into the software without requiring additional training. These resources are included in the GitHub repository (see Code Availability).

### 2.5 Overview

The complete workflow is illustrated in Fig. 1. Upon loading an EEG recording, PyHFO 2.0 operates through three primary stages: HFO detection, deep learning (DL)-based HFO classification, and results annotation. The details of each critical stage are elaborated upon in the subsequent sections.

**FIGURE 1.**
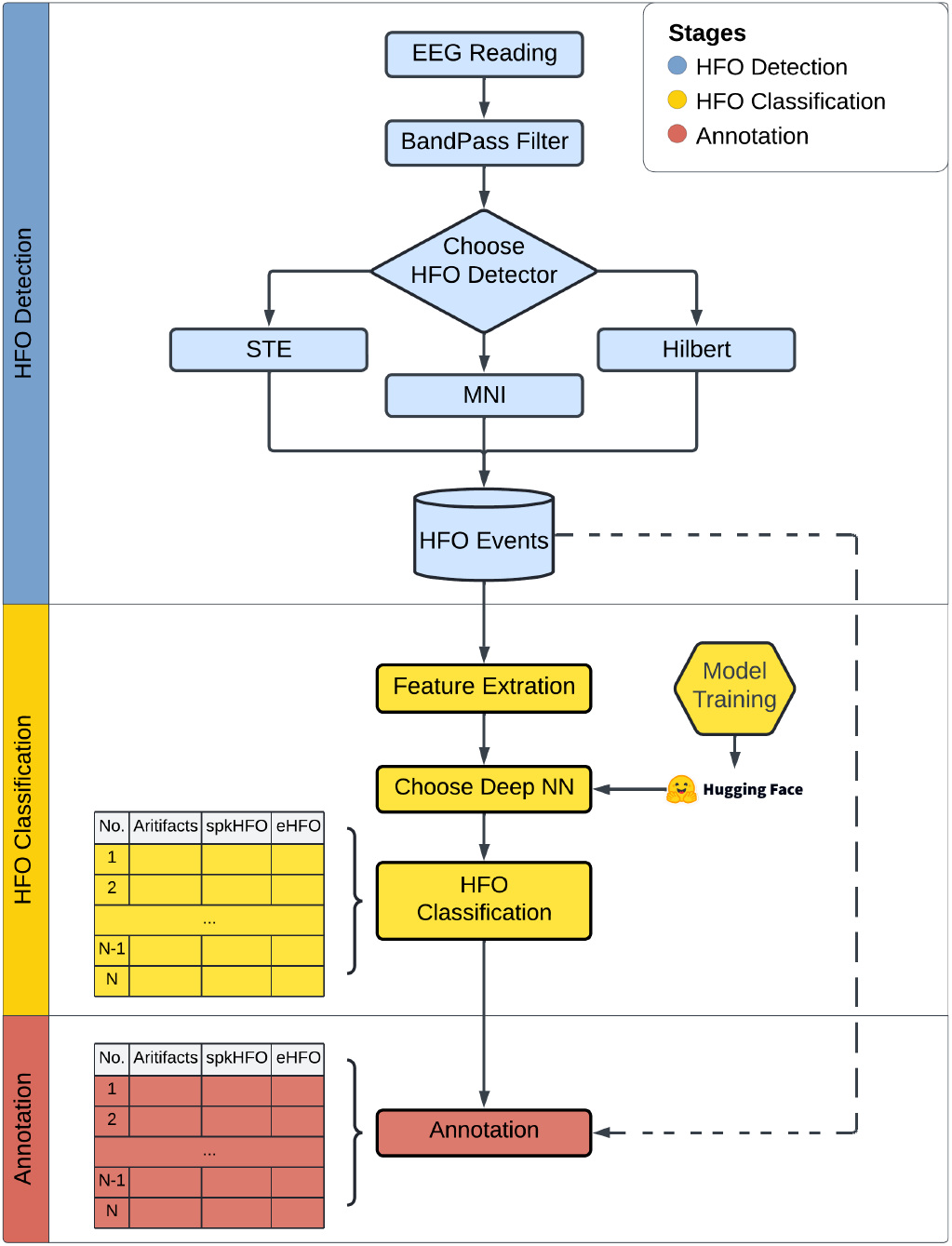
Overview of the Workflow. This flowchart illustrates the three primary stages of our study: HFO detection, deep learning classification, and annotation. The workflow significantly streamlines the efficiency of HFO analysis. Specifically, HFO detectors identify HFO events from EEG recordings and return the start and end timestamps of the detected events. The EEG signal of each detected events are sent to pre-trained deep learning models hosted on the Hugging Face Hub for HFO classification. Finally, the results can be reviewed and modified using the annotation functionality.

## 3 RESULTS

### 3.1 New Features in PyHFO 2.0

PyHFO 2.0 builds on the existing PyHFO framework with three major enhancements. First, it integrates the Hilbert (Crépon et al. 2009) method, a widely adopted HFO detection technique, thereby expanding the platform’s range of detection algorithms. Second, it introduces a new deep learning model specifically designed to detect epileptogenic HFOs (eHFOs) (Zhang et al. 2022b) with greater accuracy, and integrates all three classification models (artifact, spkHFO, and eHFO) into the Hugging Face ecosystem for automated download and streamlined model management. Third, an annotation feature has been implemented that allows users to interactively review, verify, and refine detection and classification results. A detailed verification of the newly added Hilbert detector and the eHFO classification model is provided in Clinical Validation. In the sections that follow, we offer a comprehensive guide to using PyHFO 2.0. Beyond covering these newly added features, we detail all core functionalities of the platform, equipping users with the knowledge and practical steps needed to make full use of its capabilities.

### 3.2 EEG Loading and Display

Fig. 2 illustrates the graphical user interface (GUI) of PyHFO 2.0. The “Open File” button (see arrow 1 in Fig. 2) allows users to load an EDF file containing multi-channel EEG data, which is then displayed in the “Waveform Display” panel. Basic information about the loaded EDF file, such as the file name, sampling frequency, number of channels, and recording length, is shown in the “Recording Information” viewing panel. Within the waveform panel, a vertical scroll bar enables navigation across channels, while a horizontal scroll bar facilitates navigation through time. Users can adjust the “Number of Channels to Display” and “Display Time Window” fields (arrows 2 and 3 in Fig. 2) to specify how many channels and how much time are visible in the waveform panel. Additionally, the “Display Time Window Increment” field (arrow 4 in Fig. 2) allows users to control the speed of navigation through time by adjusting the step size for the horizontal scroll bar. By pressing the “Update Plot” button, the display is dynamically refreshed to reflect the specified parameters.

**FIGURE 2.**
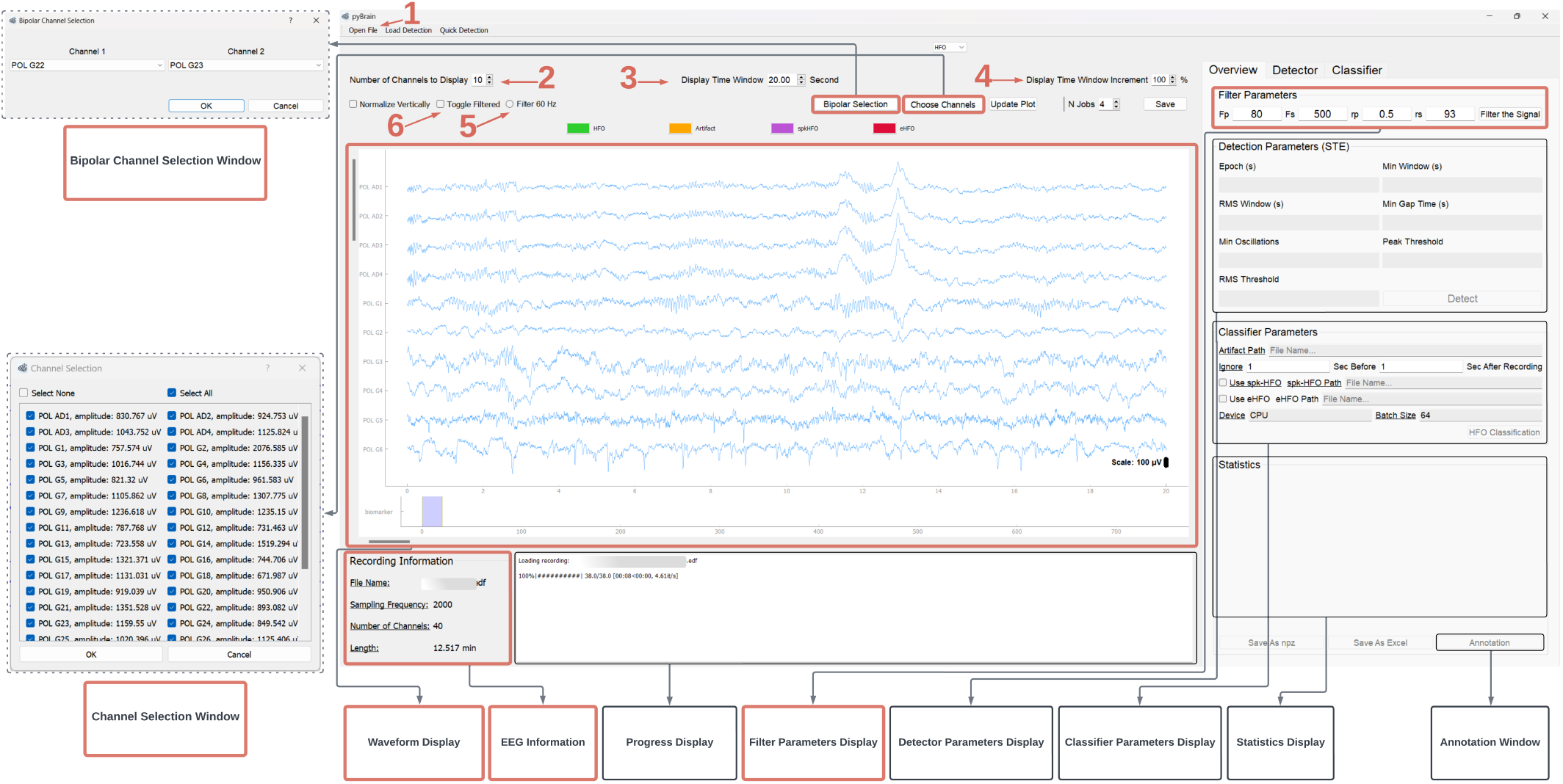
EEG loading and display. EEG loading and display functionalities are highlighting in red boxes. Arrow 1 indicates the “Open File” button, which loads an EDF file into the “Waveform Display” panel. The “Recording Information” panel displays basic information about the loaded file. A vertical scroll bar lets users navigate across channels, and a horizontal scroll bar supports time-based exploration of the EEG data. Arrows 2 and 3 mark the fields for selecting how many channels and how much time are displayed, while arrow 4 adjusts the time scroll increment. The “Choose Channels” button opens a window for selecting specific channels to view, and the “Bipolar Selection” button opens a window allowing users to form bipolar-referenced signals between any two channels. In the “Filter Parameters” panel, users can configure bandpass filter parameters. Separately, the 60 Hz interference option (arrow 5) allows removal of power-line noise. Finally, arrow 6 points to the “Toggle Filtered” checkbox, enabling users to switch between raw and filtered signals for comparison.

The “Choose Channels” button provides users with the option to select specific channels for display. When clicked, a pop-up window opens, prompting users to indicate which channels to plot. The “Bipolar Selection” button allows users to create bipolar-referenced signals by selecting pairs of channels. Clicking this button opens a popup window in which users can define and add bipolar signals according to their preferences.

To filter the signal to a desired frequency band, users can specify the filtering parameters in the “Filter Parameters” panel and then click on the “Filter the Signal” button. If the “Filter 60 Hz” radio button (arrow 5 in Fig. 2) is selected, the 60 Hz power line harmonics will also be filtered out. A Chebyshev Type II filter (implemented using SciPy) is applied for this process. Once filtering is complete, the waveform panel is automatically updated to display the filtered signal. Users can easily toggle between the raw and filtered signal by selecting or deselecting the “Toggle Filtered” checkbox (arrow 6 in Fig. 2).

### 3.3 HFO Detection

PyHFO 2.0 supports the detection of multiple biomarkers. Here, we focus on the detection of HFOs, reserving discussion of other biomarkers for later sections. PyHFO 2.0 includes three HFO detectors: STE, MNI, and Hilbert. Users can select their preferred detector in the ‘Detector’ tab and specify parameter values in the corresponding detector’s tab (see in Fig. 3). After setting these parameters, clicking the “Save” button updates the chosen parameters and displays them in the “Detector Parameters Display” panel. When the “Detect” button is clicked, the selected detector performs the detection process. The results, including the number of detected HFOs, are displayed in the “Statistics Display” panel. In addition, each detected HFO is marked at its corresponding time and channel in the waveform panel.

**FIGURE 3.**
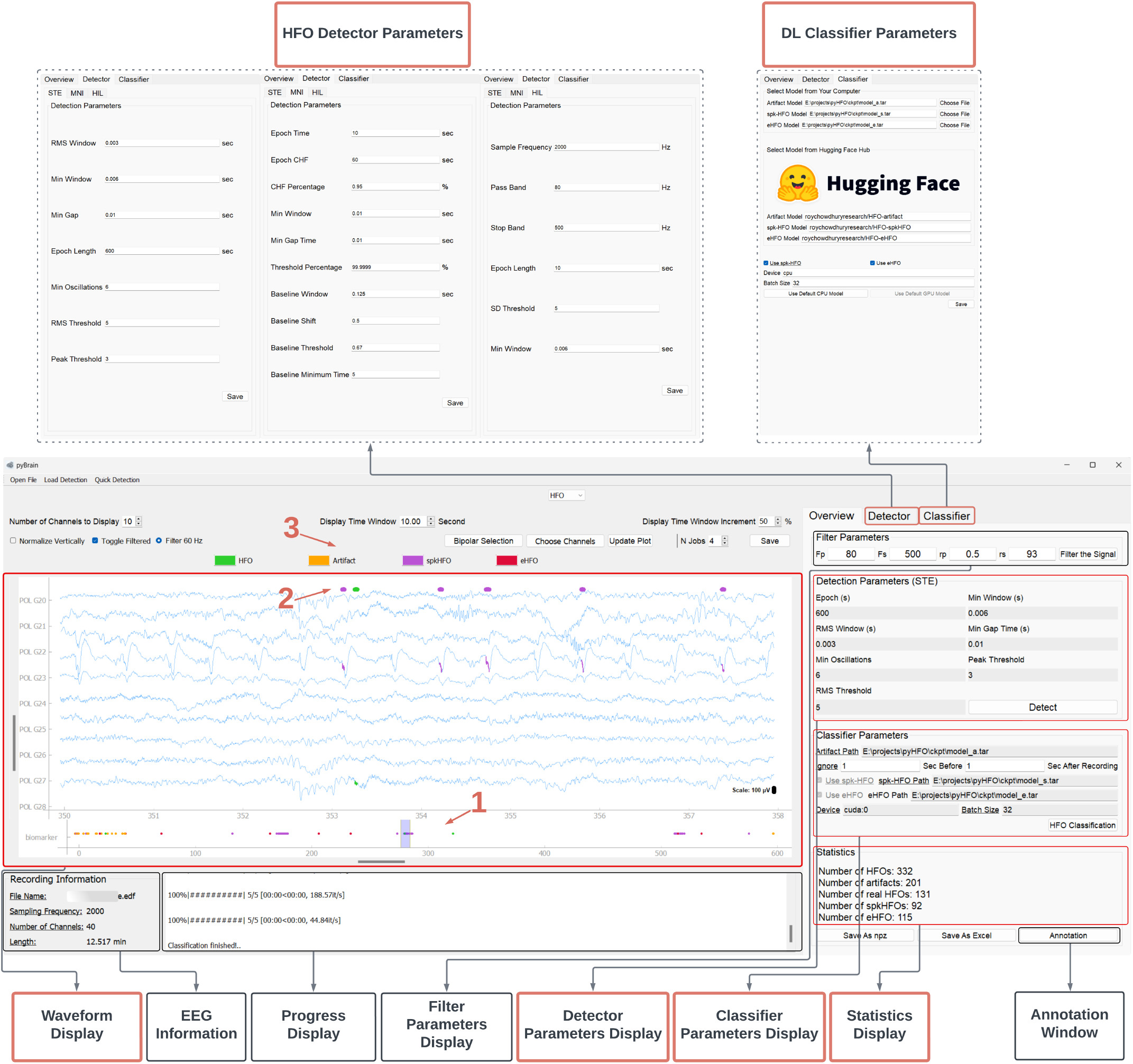
HFO event detection and classification. Key features (highlighted in red boxes) include the visualization of both HFO detection and classification results. Users can view the counts for each event category: total detected HFOs, HFOs classified as artifacts, real HFOs (non-artifacts), spkHFOs, and eHFOs. Each individual event is also overlaid with a distinct color on the signal traces in the waveform display panel (indicated by arrows 1–3). These features provide users with a clear and intuitive visualization of the HFO events.

Below the waveform panel, an event display (arrow 1 in Fig. 3) assists with navigation. The event display spans the entire EEG recording, highlighting all detected HFO occurrences within the currently displayed channels. This feature helps users align the horizontal scroll bar to quickly locate events, making exploration of detected signals more efficient.

### 3.4 Biomarker Classification

PyHFO 2.0 provides a deep learning-based HFO classification function, utilizing advanced models to ensure high accuracy and reliability. We adopted the same artifact and spkHFO classifier design as presented in a previous study, along with the eHFO classifier, given their strong performance compared to expert annotations. To ensure seamless integration and accessibility, we have uploaded our model checkpoint to the Hugging Face platform, allowing users to easily download the pretrained model directly. PyHFO 2.0 leverages the Hugging Face API to run the inference process, streamlining the user experience. This integration simplifies the setup process and enables users to take advantage of state-of-the-art models with minimal effort. In addition, if users wish to train and deploy their own classification models in the future, the process remains standardized. By adhering to Hugging Face’s framework, users can upload their custom-trained models to the platform and integrate them into PyHFO 2.0 following the same procedure.

To use the classifier, users can navigate to the “Classifier” tab and choose between two options for model loading: Option 1, specify the model path stored locally on the machine; or Option 2, specify the name of the model card stored on Hugging Face (see arrows 1–3 in Fig. 4). Users can then choose whether to run the model on a CPU or GPU (if available) and set the batch size. For convenience, two buttons, “Use Default CPU Model” and “Use Default GPU Model,” are provided for quick parameter configuration. After clicking the “Save” button, the selected parameters are displayed in the “Classifier Parameter Display” panel.

**FIGURE 4.**
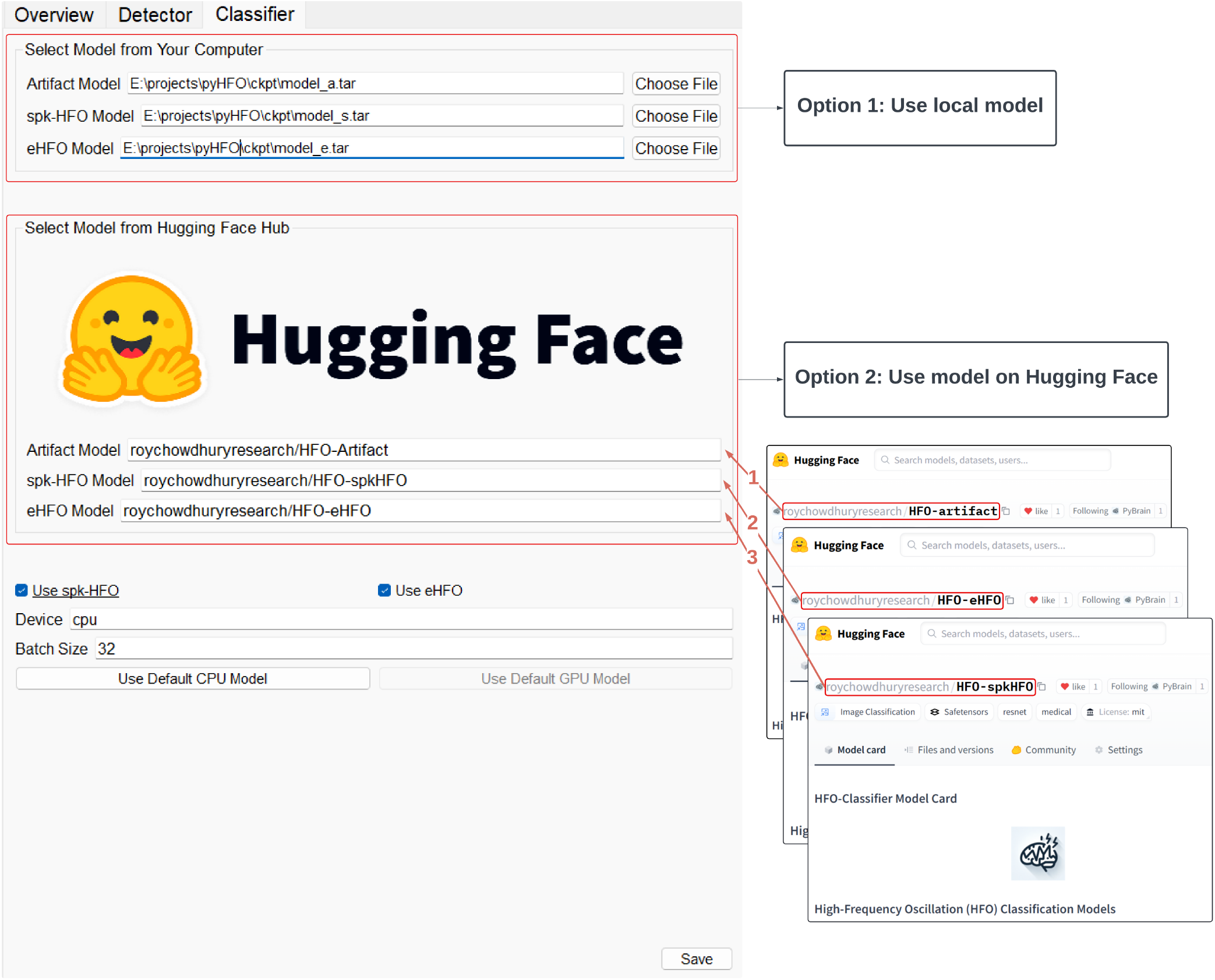
Deep Learning Classifier Parameters Setup Window. Users can specify the model path using one of two options: (1) select an existing model from the local machine, or (2) provide the model card name from the Hugging Face Hub (arrow 1-3). If users do not wish to use their own models, they can simply click on the “Use Default CPU Model” button or the “Use Default GPU Model” button (if a GPU is available), and all default parameters will be automatically loaded.

When the “HFO Classification” button is clicked, if the user selects Option 2 for model loading, the pre-trained model is automatically downloaded (stored in the local Hugging Face cache directory by default), and the classifier begins its inference process. The results, including the number of artifacts, the number of spikes, and the number of real HFOs, are displayed in the “Statistics Display” panel. Additionally, each detected HFO in the waveform panel is updated with a color code corresponding to its classified type: orange for artifacts, purple for spkHFOs, and green for HFOs, as shown in Fig. 3.

### 3.5 Result Export and Import

Prediction results can be saved in npz (NumPy) file format or Excel format using the “Save As npz” or “Save As Excel” options, respectively. Previous run results can also be imported via the “Load Detection” button, allowing users to select a previously saved npz or Excel file.

### 3.6 Event Annotation

PyHFO 2.0 also provides an annotation window, which can be accessed by clicking the “Annotation” button. This feature opens in a pop-up interface that allows users to review each detected HFO in detail. For every event, the interface displays four signal views: the raw signal, the filtered signal, a time-frequency plot (with a user-defined frequency range), and an FFT plot (also with a user-defined frequency range), all centered on the midpoint of the HFO duration (see highlighted boxes a-d Fig. 5). Both time and frequency parameters can be adjusted for more precise visualization. The “Duration Selection” drop-down menu (arrow 4 in Fig. 5) allows users to set the signal window to 1 second, 0.5 seconds or 0.25 seconds. The “Frequency Controls” section (highlighted box e in Fig. 5) lets users modify the frequency display range for both the time-frequency and FFT plots. In the “Event Information” panel (highlight box e in Fig. 5), users can view key details about the current HFO, including its channel, duration, start and end times, and the classifier predicted type (DL Suggestion). To annotate the event, users may select a new event type from the “Event Type” drop-down menu (arrow 3 in Fig. 5) and click the “Accept” button. Navigation between detected HFOs is facilitated by the “Previous” and “Next” buttons (arrow 1 in Fig. 5) and the “Event Selection” drop-down menu (arrow 2 in Fig. 5).

**FIGURE 5.**
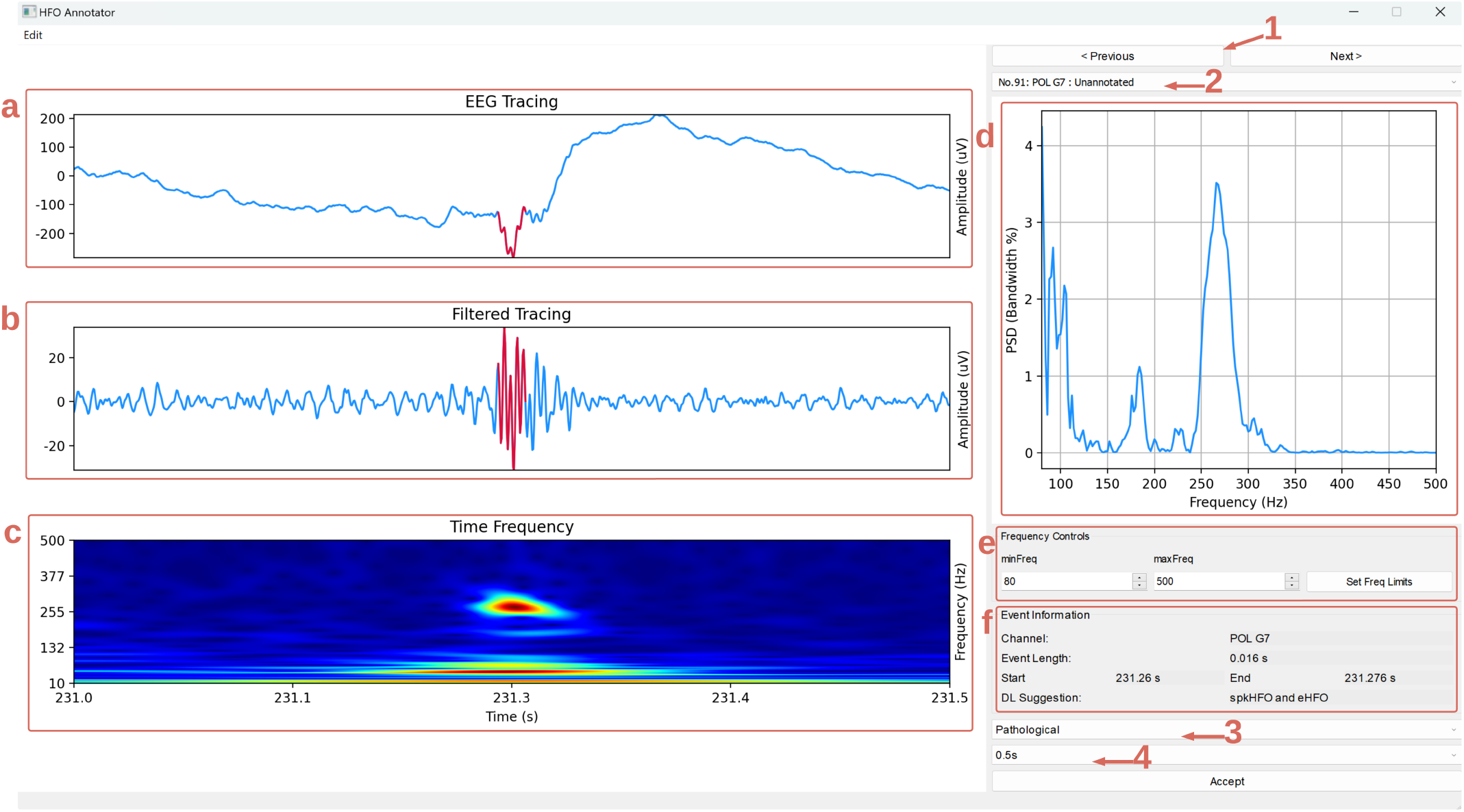
Annotation window. Screenshot of the Annotation interface in PyHFO 2.0. The raw signal, filtered signal, time-frequency plot, and FFT plot (highlighted boxes a–d) show a signal segment centered on the midpoint of the detected HFO. The “Duration Selection” control (arrow 4) and the “Frequency Controls” (highlighted box e) enable users to refine the displayed time and frequency ranges for a more detailed view of each event. Arrows 1 and 2 indicate the “Previous”, “Next”, and “Event Selection” controls for navigating among HFO events. Arrow 3 points to the “Event Type” drop-down menu, which allows users to update annotations. Additionally, box e includes the “Event Information” panel, displaying details such as the HFO’s channel, duration, start and end times, and classifier-predicted type.

### 3.7 Quick Detection

PyHFO 2.0 offers a streamlined workflow that allows users to progress from loading an EDF file to exporting HFO results within a single interface. By clicking the “Quick Detection” button, users can configure all necessary settings through a single pop-up window (Fig. 6). The process consists of the following steps:

**FIGURE 6.**
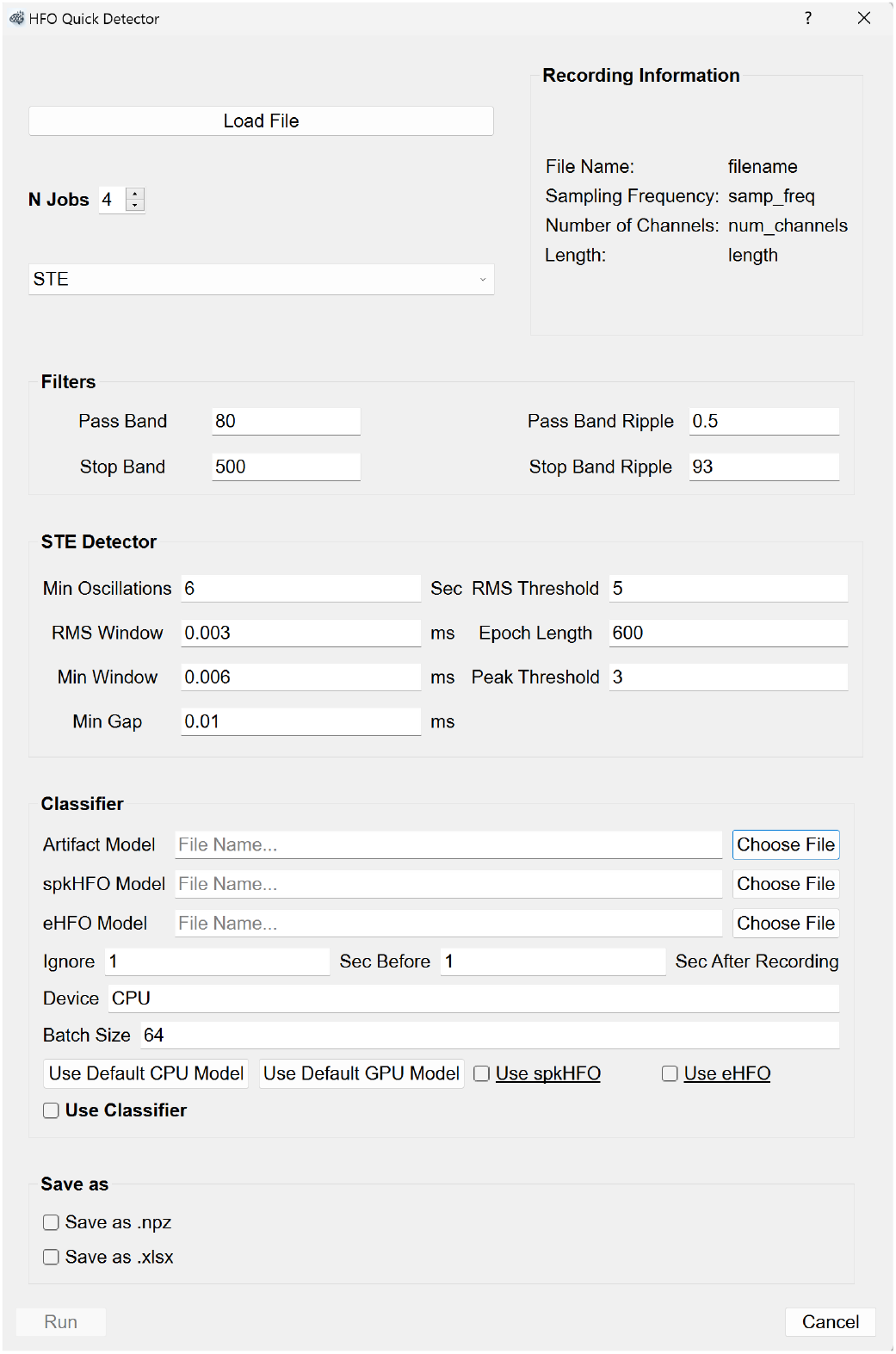
Quick Detector window. This streamlined interface allows users to configure the entire detection and classification workflow within a single pop-up window. Users can select an EDF file, adjust filtering parameters, choose a detection method, and optionally enable classification and result export. Once all settings are defined, clicking the “Run” button sequentially executes the chosen modules, generating the final results.

- Select the EDF file.
- Configure filter parameters.
- Choose a detection method and set detector parameters.
- (Optional) Enable classification, select model checkpoints, and configure classification parameters.
- (Optional) Enable result export and select the desired file format.

Once all parameters are set, clicking the “Run” button sequentially executes the chosen modules, processing the EEG data, and generating the final results.

### 3.8 Deploying Deep Learning Models

PyHFO 2.0 includes three built-in deep learning models for artifact detection, spkHFO classification, and eHFO classification. To promote flexibility and research extensibility, the platform enables users to replace these models with their own. This section outlines the standardized steps for training and integrating compatible models and demonstrates the integration process using an existing eHFO classifier.

#### 3.8.1 Model Training Requirements and Input Format

Users are free to define their own model architecture and training datasets, but to ensure compatibility, the model input format must follow the standardized feature extraction process implemented in the software. Each detected HFO event is represented by a time-frequency plot derived from a two-second EEG segment centered at the midpoint of the event. A wavelet transform is applied to obtain the timefrequency representation, covering a frequency range of 10–500 Hz sampled at 224 frequency points. To focus on the event itself, the outmost 0.5 seconds before and 0.5 seconds after the HFO event are removed from the original two-second segment, leaving a final time-frequency representation spanning 224 frequency bins across a one-second duration, with the HFO event centered. This representation is then resized to a 224 × 224 image, which serves as the input of the model. Users must ensure that their training data aligns with this format to integrate seamlessly into the classification pipeline.

#### 3.8.2 Model Integration Pathways

Once a model is trained, it can be integrated through two methods. The first option is local integration, where the trained model is saved in a PyTorch compatible format, such as `.pt` or `.pth`, and placed in the `ckpt` folder within the software directory. The second option is the integration of the Hugging Face Hub, which provides a cloud-based model hosting approach. Users can either train a model using the Hugging Face API, for which a sample training script (`train_example.py`) is provided on GitHub, or migrate an existing trained model by defining a Hugging Face model class and transferring the model weights. A migration script (`migrate_example.py`) is available to assist in this process.

By following these steps, users can replace any of the default classifiers with their own models, enabling a flexible and extensible classification framework. In our implementation, the eHFO model was migrated using `migrate_example.py` while the artifact and spkHFO models were updated with new data using the `train_example.py` script.

#### 3.8.3 Demonstration: eHFO Model Distillation and Deployment

To demonstrate how an existing deep learning model can be integrated into the PyHFO 2.0 framework, we selected the publicly available eHFO classifier as a case study. In the original implementation, a 1000 ms EEG segment centered around the HFO event midpoint was extracted and transformed into three distinct image representations: a time-frequency plot (via Gabor wavelets from 10 to 500 Hz), a tracing plot (time-domain waveform mapped onto a 2000×2000 image), and an amplitude-coding plot (where pixel intensity reflected signal amplitude over time). These three images were resized to 224×224 and concatenated to form a multi-channel input for classification.

To align this model with our standardized pipeline and reduce computational overhead, we performed a model distillation. We retained the original architecture but limited the input to a single time-frequency plot generated under the same frequency and time window (10–500 Hz over 1000 ms). This time-frequency representation was duplicated across the channel dimension to yield a 3×224×224 input, ensuring compatibility with the expected input shape of the original model.

We used the Open-iEEG dataset (Zhang et al. 2024a), which includes recordings from 185 epilepsy patients and 686,410 HFO events detected by the STE and MNI detectors, for distillation. After removing artifacts using the PyHFO artifact classification model, 332,409 valid events remained. Predictions from the original eHFO model were used as pseudo-labels to train the distilled version. This approach maintained high predictive accuracy while eliminating reliance on multiple image modalities and reducing computational cost.

Since the distilled eHFO model relies solely on the time-frequency plot input, it can be seamlessly integrated into the PyHFO platform by incorporating the relevant model definition and loading the corresponding checkpoints. The remaining procedures align with those of existing classification models, showcasing the ease of introducing new classification methods.

### 3.9 Verification of the HFO detector

To verify the newly integrated Hilbert detector in PyHFO 2.0, we followed the same validation procedure as in PyHFO by comparing the detection results with those of RIPPLELAB on three datasets (UCLA, Zurich, and Rodent), using the parameters listed in Table S2. Table S1 presents the number of HFOs detected by all PyHFO 2.0 detectors: STE, MNI, and Hilbert in two experimental settings. In the ‘Hybrid’ setting, RIPPLELAB performed data reading and bandpass filtering, while PyHFO detection was handled by PyHFO 2.0. In the “PyHFO 2.0” setting, all data reading, bandpass filtering, and detection were performed exclusively by PyHFO 2.0. Events are deemed exactly the same if they overlap by 100%, while rows 90% overlap and 50% overlap indicate how many events align for at least 90% or 50% of their intervals, respectively. Overall, these findings confirm that the Hilbert detector in PyHFO 2.0 closely aligns with RIPPLELAB, mirroring the strong agreement also observed for the previously implemented STE and MNI detectors.

**TABLE 1.**
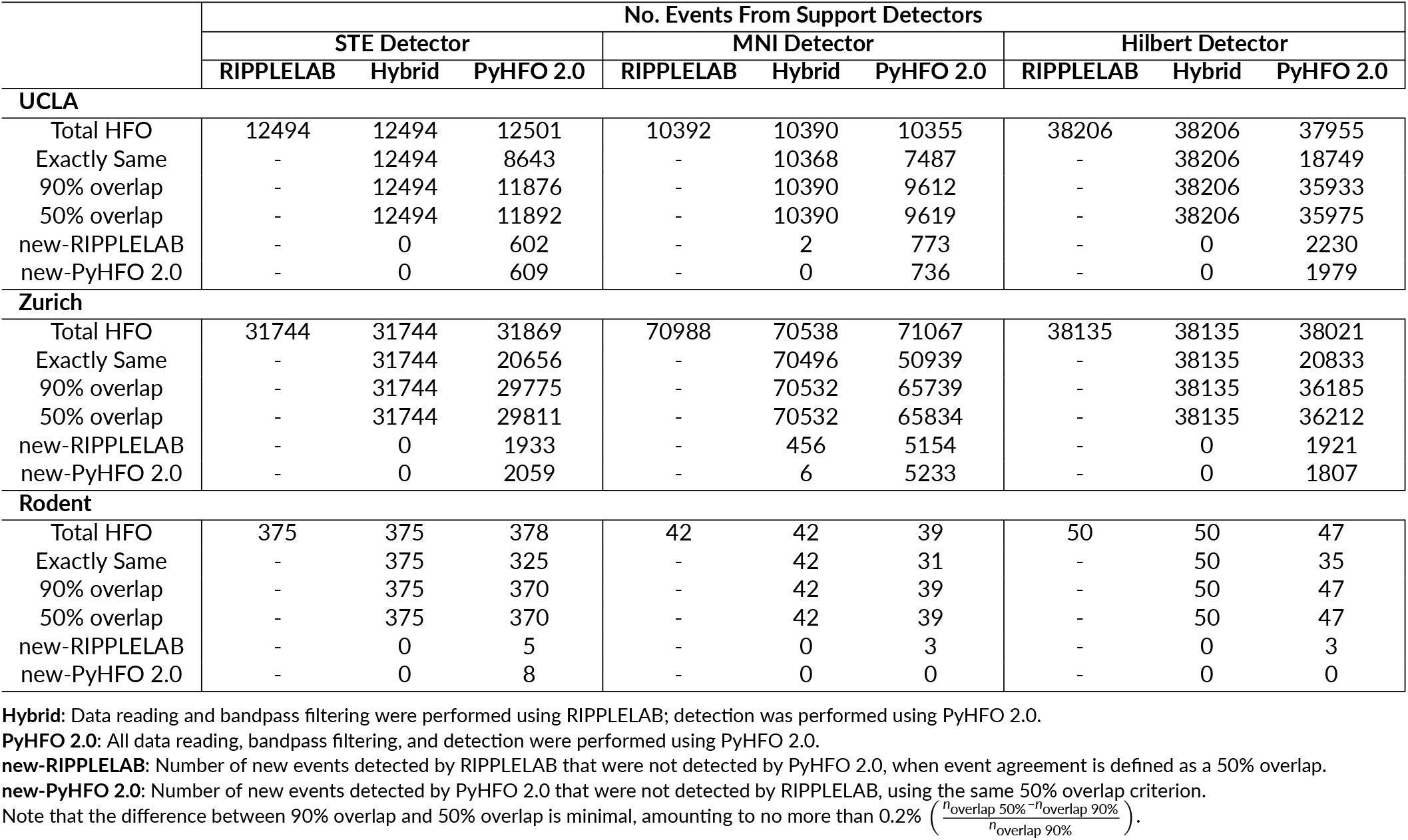
Event comparison of differences between RIPPLELAB and PyHFO 2.0 implementations in UCLA, Zurich, and Rodent datasets.

**TABLE 2.** Detailed parameters of the Hilbert-based HFO detector used for the UCLA, Zurich, and Rodent datasets.

|  | UCLA | Zurich | Rodent |
| --- | --- | --- | --- |
| filter_freq (Hz) | [80, 500] | [80, 500] | [80, 500] |
| sd_thres (in SD) | 5 | 5 | 5 |
| min_window (s) | 10*1e-3 | 10*1e-3 | 10*1e-3 |
| epoch_len (s) | 3600 | 3600 | 3600 |

filter\_freq: Frequency band for bandpass filtering.
sd\_thres: Detection threshold in units of standard deviation.
min\_window: Minimum duration for a candidate HFO to be considered valid.
epoch\_len: Length of each processing epoch in seconds.

### 3.10 Clinical Validation of DL Models

#### 3.10.1 eHFO Model Deployment Evaluation

We evaluated the performance of the distilled eHFO model by comparing it against the original eHFO model on the UCLA dataset. The goal of this evaluation is to verify that the distillation process preserves the predictive performance of the original model while significantly reducing computational overhead. Specifically, we treated the predictions of the original eHFO model as pseudo-labels and assessed how well the distilled model replicates these predictions. Accuracy was used as the evaluation metric. Our results show that the distilled eHFO model closely aligns with the original model, achieving an accuracy of 0.86.

In addition, following the methodology of Zhang et al. (2024b), we computed the computational complexity of both models. The original eHFO model requires 1.822 G MACs, while the distilled version requires only 486.95 M MACs, representing a 73 % reduction in computational cost due to distillation.

#### 3.10.2 Model Generalization on HilbertBased HFO Detection

To evaluate the generalizability of artifact and spike classification models on HFO events detected using the Hilbert transform-based detection method, we collected expert annotations from five patients in the UCLA dataset. Each detected event was independently labeled by board-certified epileptologists (AD, SK) into one of three categories: artifact, real HFO without spike, or spkHFO. Specifically, the annotated dataset includes 175 artifact events, 1366 real HFOs, and 2025 spkHFOs. Model performance was assessed by comparing model predictions with expert labels. The artifact classification model was evaluated on the full set of annotated events, treating both real HFOs and spkHFOs as non-artifactual. In contrast, the spike and eHFO classification models were evaluated only on the subset of non-artifactual events, aiming to distinguish between real HFOs and spkHFOs. The artifact classification model achieved an accuracy of 0.964, while the spike classification model reached an accuracy of 0.905.

#### 3.10.3 Seizure Outcome Prediction

Finally, we conducted a clinical evaluation of the three trained detectors and the deep learning-based HFO classification model by assessing their ability to predict surgical outcomes using the derived biomarkers. Specifically, we used the UCLA dataset as the validation cohort, which includes 15 patients with documented surgical outcomes and corresponding EEG recordings annotated by channel-level resection status. The objective of this evaluation was to build a logistic regression model using the resection ratio as a predictive feature for postoperative seizure outcomes (seizure-free vs. not seizure-free). We report the predictive performance of each biomarker in terms of the area under the ROC curve (AUC), as summarized in (Fig. 7)

**FIGURE 7.**
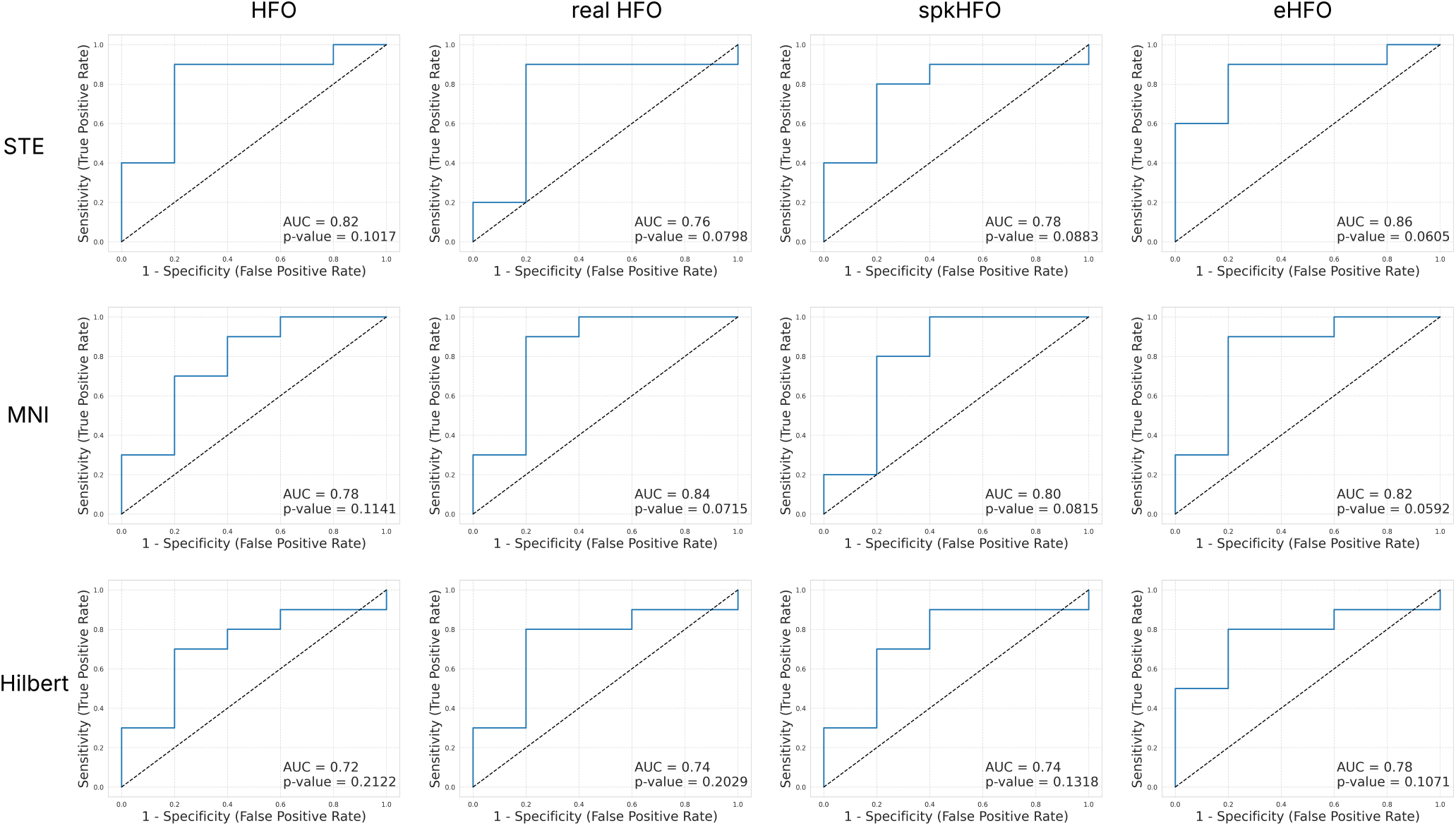
ROC curves. We constructed postoperative seizure outcome prediction models using event resection ratio derived from 10-min EEG data (n = 15). Each subfigure illustrates the classification performance for surgical outcomes using area under the ROC curve (AUC) as the evaluation metric, with p-values indicating the statistical significance of the relationship between resection ratio and seizure freedom. Rows correspond to events detected by different detectors: STE (top), MNI (middle), and Hilbert (bottom). Columns represent different event types: (1) HFO events, (2) real HFOs identified using the artifact classification model, (3) real spkHFOs identified using both artifact and spike classification models, and (4) real eHFOs identified using the artifact, spike, and eHFO classification models.

## 4 DISCUSSION

In this work, we introduced PyHFO 2.0, a user-friendly open-source platform that builds on our original PyHFO framework by integrating modern deep learning methods and incorporating additional HFO detection algorithms. PyHFO 2.0 streamlines the entire workflow—from EEG loading and filtering to HFO detection, classification, and annotation— through an intuitive graphical interface. By offering the Hilbert detection method, a dedicated eHFO classification model, and seamless integration with the Hugging Face ecosystem, PyHFO 2.0 simplifies the use of advanced machine learning models for both researchers and clinicians. The annotation window further enhances usability by allowing interactive review and refinement of detection results, thereby improving the overall reliability of HFO analysis. While PyHFO 2.0 significantly improves upon the functionality and accessibility of its predecessor, several limitations must be acknowledged:

### 4.1 Data Format Compatibility

PyHFO 2.0 currently supports reading EEG data only in the EDF and BrainVision format. This limitation may hinder adoption where other standard EEG file types (e.g., BioSemi’s .bdf) are more prevalent, requiring users to convert data before analysis.

### 4.2 Dependence on Training Data

The performance of the deep learning-based classification models relies heavily on the representativeness and quality of the training data. Users with highly specialized or underrepresented EEG datasets may need to fine-tune or retrain these models to achieve optimal performance. Future efforts could involve expanding the model’s training datasets or incorporating transfer learning techniques to better handle diverse clinical and research use cases.

### 4.3 Focus on HFOs

Although PyHFO 2.0 was developed primarily for HFO detection and classification, the platform could be extended to identify additional neurophysiological biomarkers. Implementing modules for other events, such as epileptiform discharges or spindles, broadens its applicability and potentially benefits a wider range of studies in clinical neurophysiology.

### 4.4 Image-Based Representation of HFOs

The current deep learning classification pipeline assumes that biomarkers to be classified can be represented in an image-like format (for example, through time-frequency spectrograms). While this approach works well for many HFO-related events, it may be less suitable for biomarkers that do not neatly lend themselves to two-dimensional representations. Additional preprocessing steps or alternative pipeline designs may be necessary to support a broader range of biomarker signals.

### 4.5 Future Directions

Moving forward, we plan to enhance the waveform display functionality in both the main GUI and the annotation window by introducing interactive cursors, zooming features, and improved navigation tools, allowing users to more easily inspect specific signal segments. We also intend to expand the platform to detect and classify additional neurophysiological biomarkers, with spindle detection as a priority target for future implementation. Furthermore, integrating newer deep learning techniques, such as transfer learning approaches, could improve classification performance across various data sources.

Overall, PyHFO 2.0 represents a significant advancement in bridging the gap between deep learning innovations and clinical neuroscience. By providing a flexible, intuitive, and open-source framework for HFO detection, classification, and annotation, PyHFO 2.0 equips researchers and clinicians with a powerful tool to accelerate and enrich biomarker analysis.

## Data Availability

All data produced in the present study are available upon reasonable request to the authors

## Abbreviations

EEG: electroencephalography;
HFO: high-frequency oscillation;
iEEG: intracranial electroencephalography;
STE: short-term energy;
MNI: Montreal Neurological Institute;
FFT: fast Fourier transform;
DL: deep learning;
eHFO: epileptogenic high-frequency oscillation;
spkHFO: spike-associated highfrequencyoscillation;
GUI: graphical user interface.

